# Genomic and Patient Epidemiology of *Streptococcus dysgalactiae* Subspecies *equisimilis* in Houston, Texas

**DOI:** 10.1101/2025.11.17.25340401

**Authors:** Lydia Pouga, Stephen B Beres, Randall J Olsen, S Wesley Long, Edward A. Graviss, James M Musser

**Author notes:** Corresponding author: James M. Musser, M.D., Ph.D. Lydia Pouga and Stephen B. Beres contributed equally to this work. Author order was by mutual agreement of the authors.

## Abstract

Many countries have reported increased human infections caused by *Streptococcus dysgalactiae* subspecies *equisimilis* (SDSE) in the last 15 years. However, there is scant molecular epidemiology data for SDSE in the United States, especially at the whole genome level. To address this knowledge deficit, we studied SDSE infections in a large health care system in the Houston, Texas, metroplex, an ethnically diverse population of 7.4 million. We used Illumina whole genome sequencing to characterize 865 human isolates collected consecutively from unique patients with diverse infections in the Houston Methodist Hospital system between June 2022 and August 2024. Genomic clustering assigned the isolates to 44 distinct genetic lineages (GL). GLs had a stronger correlation with the population structure as reflected by the single nucleotide polymorphism phylogenetic tree than *emm* or multilocus sequence type. We found absolute recombination nearly twice as prevalent in SDSE as in *Streptococcus pyogenes*, its closest genetically related species. This finding may explain why in contrast to *S. pyogenes*, *emm* typing is a poor molecular marker of overall genomic relationships in SDSE. We identified significant isolate genotype–patient phenotype associations: GL01 was associated with skin or soft tissue infections, GL02 with blood and urine infections, whereas GL03 was associated with throat infections. GL02 was also associated with increased clinical severity. In the aggregate, our study provides new information about SDSE infections occurring in a large metropolitan area in the United States, and expands our genomic epidemiology understanding of SDSE, as an emerging human bacterial pathogen of increasingly recognized importance.

**IMPORTANCE:** Our study provides considerable new information about the genomic epidemiology and patient characteristics of SDSE infections in a large metropolitan area in the United States. We discovered that all abundantly occurring genetic lineages were comprised of isolates with multiple *emm* gene types and multilocus sequence types. Analyses based solely or predominately on these two commonly employed molecular epidemiologic markers obscure a detailed understanding of SDSE genetic diversity, population genomics, and may fail to reveal important disease associations. Our work highlights the need for longitudinal SDSE whole genome sequencing-based surveillance and analysis of this emerging human pathogen. Such efforts will contribute to enhanced epidemiologic understanding and patient demographics, and may aid improved diagnostics, infection control, public health strategies, and vaccine development for a pathogen that disproportionately affects older patients and patients with underlying medical conditions. These at-risk populations are currently rapidly expanding in the United States and many other high-income countries.

---

*Streptococcus dysgalactiae* subspecies *equisimilis* (SDSE) is a member of the pyogenic group of streptococci that typically expresses Lancefield group C or G carbohydrate antigens, and less commonly group A (1, 2). At the species level, it is most closely genetically related to the well-studied human pathogen, *Streptococcus pyogenes*. SDSE causes non-invasive and invasive diseases, such as pharyngitis, skin and soft-tissue infections and life-threatening infections including septicemia, necrotizing fasciitis and toxic shock syndrome (3–9). Although multiple countries have reported an increasing number of human SDSE invasive infections, data remain scant (10).

The molecular epidemiology and population genetics of SDSE is poorly understood, with few whole genome sequencing (WGS)-based investigations conducted in the United States and elsewhere. Several problems limit our understanding of SDSE molecular epidemiology, including clinical laboratory identification and classification of this pathogen. For example, SDSE can be confused with other streptococci that express Lancefield group C, G or A antigens (1, 2). *emm* gene typing, which characterizes a highly variable region of the M protein gene, is a long used molecular marker for epidemiologic studies and isolate classification of *S. pyogenes* and has been adopted for use in SDSE. Early comparative studies established a structural and functional homology between M proteins of *S. pyogenes* and Group G streptococcus (11, 12), the functional diversity and clinical implications of SDSE M protein remain poorly characterized compared to *S. pyogenes* (10). However, evidence is accumulating for SDSE that *emm* typing correlates poorly with overall genomic relationships and phylogeny. Multiple studies have noted a lack of congruency between *emm* types and clonal complexes or genetic clades (10, 13–15), consistent with this species having abundant genetic recombination capable of generating considerable genomic plasticity.

While numerous countries have reported an increasing frequency of human SDSE infections, and some have speculated at heightened disease severity, analyses combining comprehensive population WGS with rich clinical data are lacking, particularly within large metropolitan areas in the United States. To address this data deficit, we analyzed SDSE infections in patients in the Methodist Hospital system located in Houston, Texas. We sequenced the genomes of 865 SDSE isolates cultured from 865 unique patients with diverse invasive and non-invasive infections collected over two years. SDSE isolates in this large sample are genetically highly diverse. Although *emm* gene typing and multilocus sequence typing (MLST) are commonly used to classify SDSE isolates for epidemiological purposes, our data indicate that focusing solely or predominately on these molecular marker schemes obscures understanding of SDSE genomic diversity, overall population genetic relationships among isolates, and correlations between isolate genotype and patient phenotype. Taken together, the data provide new information about SDSE isolates causing human infections in a major metropolitan population center of the United States.

## RESULTS

### Patient sample

Patient isolates were presumptively identified as SDSE by MALDI-TOF in our central diagnostic microbiology laboratory, and subsequently serologically assigned to Lancefield group carbohydrate antigen using latex agglutination. Isolates were Illumina short-read whole genome sequenced, resulting in definitive identification of genus, species, and subspecies. In total, 865 SDSE isolates cultured from consecutive unique patients from June 2022 to August 2024 were investigated. This sample is a comprehensive collection of organisms recovered from patients in the large Houston Methodist healthcare system during this period. Patient demographic, case clinical, and isolate molecular genetic data are presented in Table S1.

### Sample overview

The anatomic collection sites for the 865 SDSE isolates were as follows: 604 (69.8%) isolates were cultured from throat, 108 (15%) from skin/soft tissue, 92 (10.6%) from blood, 40 (4.6%) from urine, 8 (0.9%) from lower respiratory tract, 6 (0.7%) from joint fluid, 4 (0.5%) from bone, and 3 (0.3%) from the peritoneum (Table S1).

### Patient demographics

The mean patient age was 36 years (range: 7 months to 96 yr). The cohort was comprised of 136 pediatric patients (< 18 yr, 16%), 617 adult patients (18–64 yr, 71%), and 112 senior patients (≥ 65 yr, 13%). Modestly more patients, 57% (*n* = 490) were female than male. There was extensive racial and ethnic diversity in the 865 patients, an observation reflecting the fact that Houston is one of the most demographically diverse cities in the United States (16). Patients identifying as Black or African American were most abundant (*n* = 361, 42%), followed by White non-Hispanic (*n* = 242, 28%), Latino or Hispanic (*n* = 180, 21%), and Asian (*n* = 34, 4%). Patients from other ethnicities made up 2% (*n* = 19) of the sample. The race or ethnicity was unknown for 29 patients (3%).

### Analysis of patients by infection specimen site

To test the hypothesis that there were significant differences in clinical characteristics across infection sites, we performed an ANOVA for age, and Chi-square for categorical clinical parameters (Table S2). Patients differed markedly across infection specimen types, with respect to age (*P* < 0.001), gender (*P* < 0.001), presence of underlying diseases (*P* < 0.001), disease severity (*P* < 0.001), and poor outcome (*P* < 0.001). Patients with throat and urine infections were younger (mean age, 26 and 47 years, respectively) and predominantly female (62% and 85%, respectively), whereas patients with skin/soft tissue or blood infections were older (mean age, 55 and 64 years, respectively) and predominantly male (65% and 63%, respectively).

Almost one-third of patients with blood infections had a severe clinical presentation, as defined by admission to the ICU and/or presence of shock, whereas 1% or less of patients with other infections had a severe presentation. Patients with skin/soft tissue or blood infections, disproportionately the elderly, also had a greater burden of underlying diseases, particularly cardiovascular disease (71% and 90%, respectively) and diabetes mellitus (46% and 51%, respectively). Patients with skin/soft tissue or blood infections also had the highest proportion of poor outcomes (12% and 11%, respectively), whereas no patients with throat or urine infections had poor outcome, as defined by disability, amputation, and death.

### Conventional molecular marker-based classification

Among the 865 isolates, Lancefield group G antigen expression was detected in 500 (57.8%) isolates, group C in 356 (41.2%), and group A in 8 isolates (0.9%) (Table S1). A lone isolate was reproducibly positive for expression of both group C and G antigens (0.1%). 44 distinct *emm* types were identified (FIG 1A and Table S1). The majority of the isolates (51%) are encompassed by the seven most numerically abundant *emm* types: STC839 (*n* = 101, 12%), STG62647 (*n* = 87, 10%), STC74A (*n* = 68, 8%), STG653 (*n* = 55, 6%), STG245 (*n* = 44, 5%), STG4974 (*n* = 42, 5%), and STG485 (*n* = 40, 5%).

**FIG 1.**
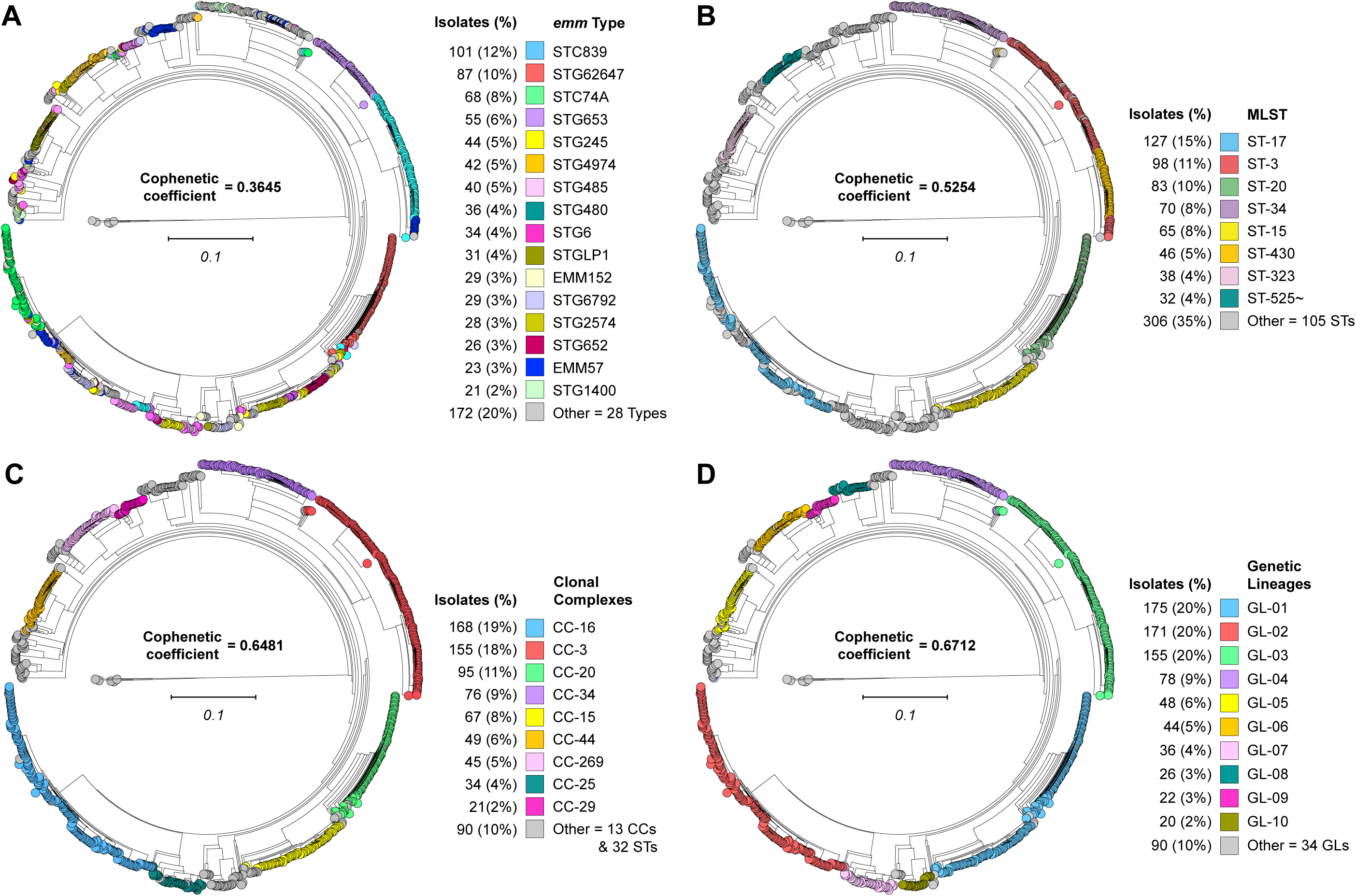
Phylogenomic relationships among SDSE isolates and correlation with molecular typing methods. Illustrated as midpoint-rooted circular cladograms are phylogenies for all 865 isolates studied, inferred by neighbor-joining based on 159,360 concatenated core chromosomal SNPs. For each typing method, groups consisting of 20 or more isolates are individually colored as per each panel index. (A) *emm* gene typing identified 44 types. (B) Multi-locus sequence typing (MLST) identified 114 STs. (C) MLST single-locus-variant analysis identified 22 clonal complexes (CCs) and 32 singleton STs. CCs were designated by the lowest numbered ST encompassed. (D) Genetic lineages (GLs) defined by hierarchical density-based spatial clustering identified 44 lineages. Correspondence of each typing method with the phylogeny was evaluated by calculation of the cophenetic correlation coefficient. GLs had the highest and *emm* typing the lowest correlation with the phylogenomic population structure.

We identified 113 MLSTs (FIG 1B and Table S1). The majority of the isolates (51%) are encompassed by the five most abundant MLSTs: ST-17 (*n* = 127, 15%), ST-3 (*n* = 98, 11%), ST-20 (*n* = 83, 10%), ST-34 (*n* = 70, 8%), and ST-15 (*n* = 65, 8%). Using the criterion of single locus variation, genetically related STs were aggregated into clonal complexes (CCs). This analysis resulted in the identification of 22 CCs and 32 singleton STs within the population (FIG 1C and Table S1). The majority of the isolates (57%) are encompassed by the four most abundant CCs: CC-16 (*n* = 168, 19%), CC-3 (*n* = 155, 18%), CC-20 (*n* = 95, 11%), and CC-34 (*n* = 76, 9%).

### Whole genome sequencing-based classification

To establish a population genomic framework for investigating isolate genotype-infection phenotype relationships among the 865 SDSE isolates, genetic lineages were delineated by hierarchical density-based spatial clustering with PopPUNK (17) (FIG S1). Isolates clustered into 44 genetic lineages (GLs) of core genomes designated GL01 to GL44 in decreasing order of abundance (FIG 1D and Table S1). Within a GL, isolates differed on average by 2,294 core SNPs, and between GLs by 18,049 core SNPs. The ten most abundant GLs accounted for 90% of the 865 isolates.

### Congruence between phylogenetic structure and classification schemes

To evaluate how well the various classification schemes (*emm*, MLST, CC, and GL) accurately capture the phylogenetic population structure, we calculated cophenetic correlation. The coefficients for the classification schemes correlation with the phylogeny decreased in the following order: GLs (0.6712), followed by CCs (0.6481), then STs (0.5254), and lastly *emm* types (0.3645). This analysis strongly demonstrates that for SDSE, GLs more accurately capture the inferred population genomic structure than *emm* typing or MLST (FIG 1). We next evaluated the similarity of the clusterings by the classification schemes against each other using the Adjusted Rand Index (ARI). The strongest similarity was observed between GLs and CCs (0.8752), whereas between GLs and STs (0.6289) or GLs and *emm* types (0.3598), similarity was weaker (Table S3). Finally, we evaluated how well classification of isolates by one scheme was predictive of classification by another and vice-versa, using the Wallace index. GLs shared the highest reciprocal predictive capacity with CCs and the lowest with *emm* type (Table S3). Taken together, these results unambiguously establish that classification by GL better captures the phylogenetic population structure than classification by *emm* typing or MLST. Thus, subsequent isolate genotype–infection phenotype association analyses are based on GL classifications.

### Recombination assessments

The poor correlation of *emm* type with the phylogenetic population structure for the SDSE isolates studied is consistent with multiple prior investigations that have all found: 1) the same *emm* type present in multiple divergent genetic backgrounds, and 2) numerically abundant genetic backgrounds to be composed of multiple different *emm* types (13, 15, 18, 19). The 10 most numerically abundant genetic lineages (GL01 to GL10), each encompassed multiple *emm* types (3 to 17) and multiple MLSTs (3 to 19). Moreover, 70% (31/44) of *emm* types were present in multiple GLs (2 to 11). These findings have been attributed to the *emm* gene being horizontally transferred among divergent genetic backgrounds at sufficient frequency to result in *emm* type being an unreliable molecular marker on epidemiologically relevant time scales (20). To address the hypothesis that the poor correlation of *emm* type with genetic background is a consequence of high level of horizontal genetic transfer and recombination in SDSE, we assessed recombination within the core genome sequences of all 865 isolates using Gubbins (21) and ClonalFrameML (22). Inferred recombination was extensive throughout the phylogeny for the 865 SDSE isolates (SDSE-865) with blocks of recombined sequence occurring all along the SDSE core genome (FIG 2A). Within the SDSE-865 core genome phylogeny, 18,184 unique recombination events were inferred (Table 1), with the start position of recombination blocks (RBs) spaced on average every 107 bps (range = 0 to 5055 bp) along the core chromosome. The metrics of recombination for the 865 SDSE core genomes are comparable to that previously determined for 595 *S. pyogenes* (Spyo-595) core genomes selected to be representative of the species’ known genetic diversity (23). Although the recombination to mutation rate ratio (ρ/θ) was nearly identical between the species, the mean length (δ) and diversity (ν) of RBs was greater in SDSE, resulting in a ∼1.5-fold greater effective recombination to mutation rate ratio in SDSE relative to *S. pyogenes*. In the SDSE core genome, a nucleotide change was determined to be 2.4 times as likely to stem from a horizontal recombination event as from a vertical single-site mutation event.

**FIG 2.**
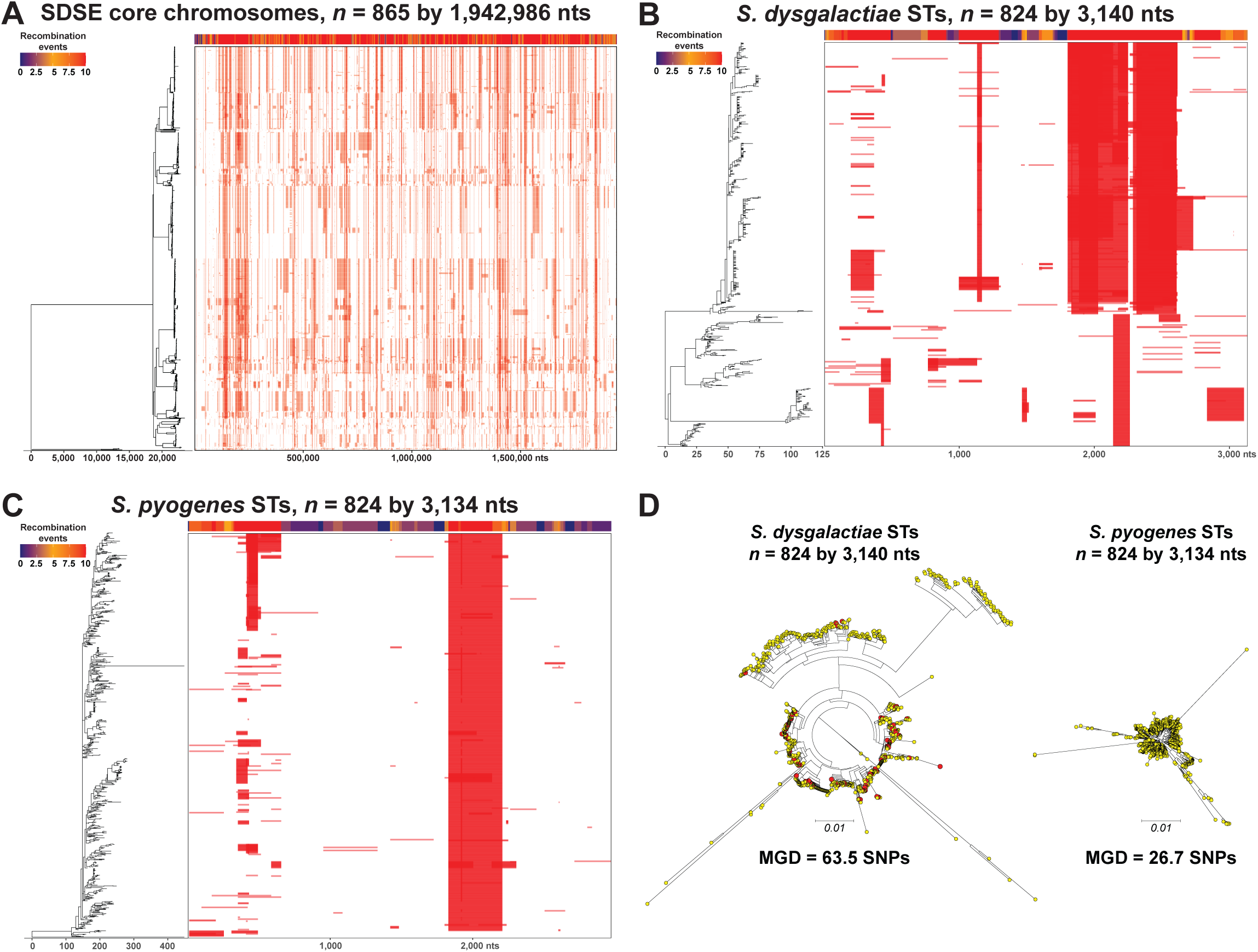
Assessment of recombination. Illustrated in panels A, B, and C are phylogenies (on the left) and recombination block diagrams (on the right) with a heatmap of recombination events (along the top) as inferred with Gubbins. (A) Recombination blocks (*n* = 18,184) in the phylogeny of 865 SDSE isolates across the 1,942,986 nt core chromosome. (B) Recombination blocks (*n* = 363) in the phylogeny of 824 *S. dysgalactiae* STs along the 3,140 nt aligned sequences. (C) Recombination blocks (*n* = 189) in the phylogeny of 824 *S. pyogenes* STs along the 3,134 nt aligned sequences. (D) Phylogenetic relationships among the 824 *S. dysgalactiae* STs and the 824 *S. pyogenes* STs inferred by neighbor-joining and illustrated as mid-point rooted circular cladograms. The phylogenetic trees are at the same scale. Shown in red in the *S. dysgalactiae* ST tree are the 61 of 824 STs overlapping with those present among the 865 SDSE isolates studied.

**TABLE 1.**
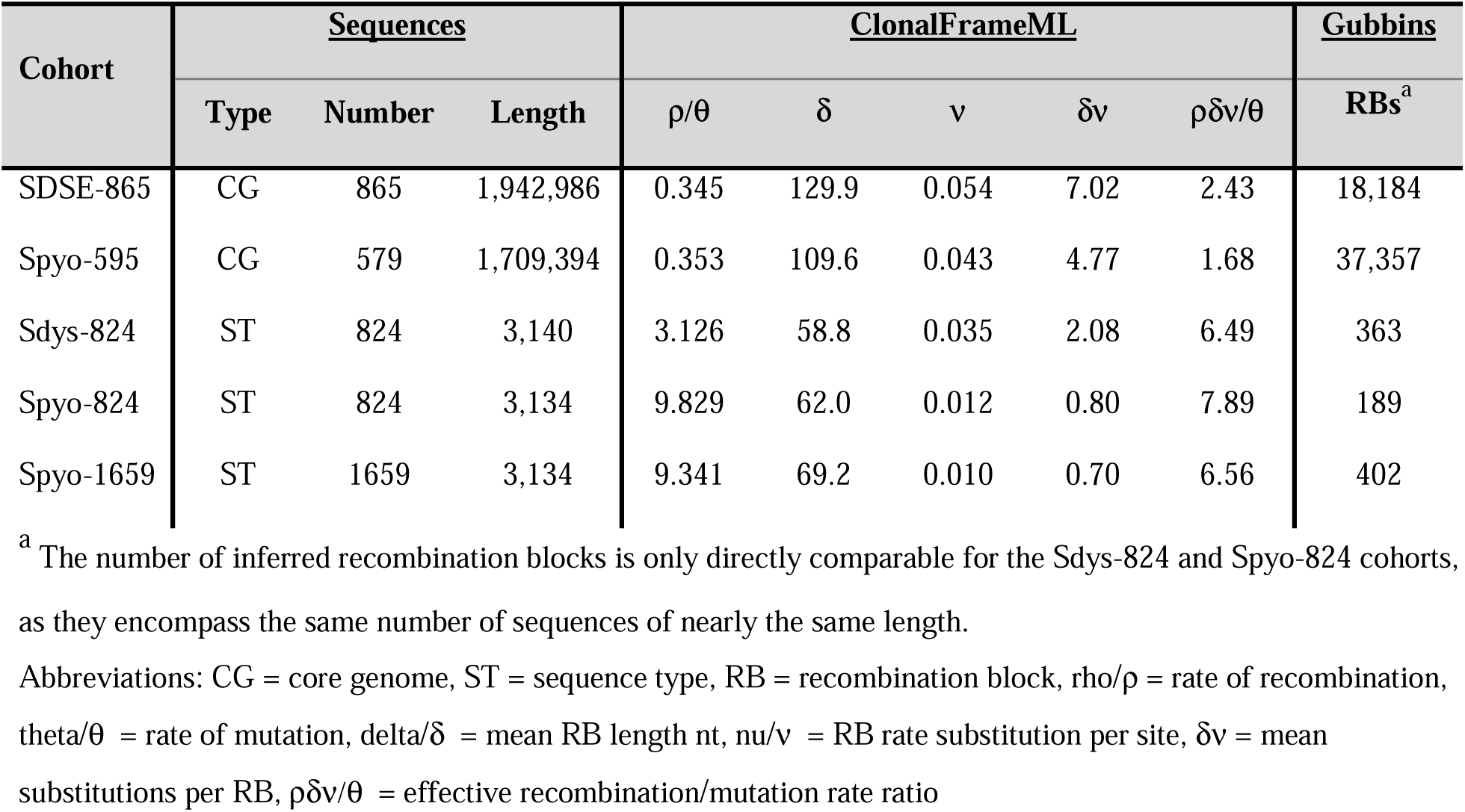
Recombination metrics.

Different bacterial species and genetic lineages within a species can exhibit substantial variation in their molecular evolutionary rates (24–27). If isolates within a species evolve at substantially different rates, then the species as a whole violates the assumption of a strict molecular clock. To test the hypothesis that different SDSE isolates may be evolving at different rates, we assessed recombination within the core genome for each of the 10 most numerically abundant GLs individually (FIG S2.1, S2.2). These 10 GLs are each composed of 20 or more isolates and account for 89.6% (775/865) of the isolates. Although there was GL-to-GL variation in the recombination metrics determined, for any metric the variation was less than two standard deviations from the mean. Moreover, as determined by Grubb’s outlier test, none of the GLs had a metric that was a significant outlier (FIG S3). Thus, despite the extensive recombination present among the core genome phylogenies, these findings support the potential for SDSE to be evolving in a more-or-less clocklike fashion.

Recombination analysis of the core genome revealed that SDSE exhibits a per-site recombination-to-mutation rate ratio (ρ/θ) that is nearly identical to that of *S. pyogenes* (Table 1). However, comparing this ratio alone does not assess the absolute frequency of effective recombination events across the broader species-wide population. To assess the absolute frequency of recombination events for both SDSE and *S. pyogenes* on a broader scale, we analyzed an equal number of MLST sequences obtained from the PubMLST database. The database contained 824 *S. dysgalactiae* STs and 1,659 *S. pyogenes* STs (as of 9/16/2025). Since PubMLST does not specify *S. dysgalactiae* at the subspecies level, we compared all 824 *S. dysgalactiae* STs (Sdys-824-ST) with a size-matched subset of *S. pyogenes* STs (Spyo-824-ST). Facilitating the cross-species comparison, six of the seven MLST alleles utilize internal fragments of the same housekeeping genes (*gki, gtr, murI, mutS, recP, and xpt*), and these genes are conserved in the same chromosomal context between the species. The resulting aligned concatenated sequences are nearly identical in length (Sdys-824-ST = 3,140 nt; Spyo-824-ST = 3,134 nt) (FIG 2B, 2C). The analysis determined that the effective recombination-to-mutation rate ratio (ρδν/θ) was only modestly lower in Sdys-824-ST (6.49) than in Spyo-824-ST (7.89), and nearly identical to the rate found in the larger Spyo-1659-ST (6.56). Crucially, despite this near equivalence in the effective rate ratio, the absolute number of inferred recombination events was nearly twice as high in the Sdys-824-ST phylogeny (*n* = 363) compared to the Spyo-824-ST phylogeny (*n* = 189). Further supporting the hypothesis that SDSE has a higher rate of evolution than *S. pyogenes*, the pairwise average genetic distance between the 824 *S. dysgalactiae* STs (63.5 SNPs) is also approximately double that between the 824 *S. pyogenes* STs (26.7 SNPs) (Fig. 2D). Taken together these findings suggest that SDSE may be evolving or diversifying at twice the rate of *S. pyogenes*.

### Association of GLs with infection sites

Previous SDSE molecular epidemiology investigations have either not attempted or have not conclusively identified isolate genotype-infection phenotype associations (1, 18, 19, 28). Having established that GL better captures genomic relationships among isolates than MLST or *emm* typing, we next tested the null hypothesis that infection site phenotype was independent of GL genotype. First, this was addressed globally including all GLs relative to all infection sites. The statistical significance of these associations was evaluated using the Fisher-Freeman-Halton exact test with 10^5^ Monte Carlo permutations to calculate the non-parametric *P*-value. These analyses identified a significant association (both *P* < 0.001), revealing that infection sites are not independent of GLs.

We next individually tested the ten most prevalent GLs for significant non-random associations with the four most prevalent anatomic sites (40 individual tests). Multiple associations, both positive and negative, were identified (Table S4). After adjusting for multiple comparisons, we identified significant positive associations between GL01 and skin/soft tissue isolates (*P* < 0.001), GL02 and blood isolates (*P* < 0.001) and urine isolates (*P* = 0.001), and GL03 and throat isolates (*P* < 0.001) (Table 2). To better assess the strength of the association between GLs and anatomical site, we calculated odds ratios. The odds of skin and soft tissue infection were more than twice as high in patients infected with GL01 isolates compared to those with non-GL01 isolates (OR = 2.338, 95% CI: 1.508–3.623, *P* < 0.001). For blood and urine infections, the odds were nearly three times higher in patients infected with GL02 isolates than in those with non-GL02 isolates (blood: OR = 2.721, 95% CI: 1.715–4.319, *P* < 0.001; urine: OR = 2.882, 95% CI: 1.495–5.555, *P* = 0.002). The highest ORs were observed for throat infections, which were almost six times more likely in patients infected with GL03 isolates compared to non-GL03 isolates (OR = 5.863, 95% CI: 3.255–10.562, *P* < 0.001).

**TABLE 2.** Chi Square analysis of genetic lineage and isolate anatomic site association.

| Genetic Lineage | Throat |  | Skin/Soft tissue |  | Blood |  | Urine |  |
| --- | --- | --- | --- | --- | --- | --- | --- | --- |
|  | Obs/Exp | P-value | Obs/Exp | P-value | Obs/Exp | P-value | Obs/Exp | P-value |
| GL01 | 0.81 | 0.00002<sup>a</sup> | 1.70 | 0.00010<sup>a</sup> | 1.13 | 0.51222 | 1.11 | 0.71556 |
| GL02 | 0.81 | 0.00003<sup>a</sup> | 0.93 | 0.72728 | 1.87 | 0.00001<sup>a</sup> | 2.03 | 0.00100<sup>a</sup> |
| GL03 | 1.31 | < 0.00001<sup>a</sup> | 0.46 | 0.00549 | 0.18 | 0.00011<sup>a</sup> | 0.14 | 0.00922 |

<sup>a</sup> significant P-value after Benjamin-Hochberg correction.

### Association of GLs with infection severity and outcome

We next tested the hypothesis that GLs were associated with infection severity or outcome (Table 3). The analysis was limited to the five largest GLs (GL0-GL05), which accounted for 73% of isolates and provided adequate sample sizes for robust association testing. As age correlated with infection severity and poor outcome, we also calculated odds ratios adjusted for patient age. Only GL02 was significantly associated with infection severity. The odds of severe disease was approximately three times higher in patients infected with GL02 than with non-GL02 isolates (OR = 3.082, 95% CI (1.553–6.114), *P* = 0.001). In multivariate logistic regression with age as a covariate, the relationship between GL02 and severe clinical presentation remained significant (OR = 2.607, 95% CI (1.278–5.320), *P* = 0.008). No other GL was associated with infection severity, and none of the five GLs tested were associated with poor outcome.

**TABLE 3.** Logistic regression analysis of genetic lineage with disease severity and poor outcome association.

| Risk | Outcome<sup>a</sup> | OR (95% CI) | *P*-value | OR adjusted for age (95% CI) | *P*-value<sup>b</sup> |
| --- | --- | --- | --- | --- | --- |
| **GL01** | Severity | 1.331 (0.614 – 2.885) | 0.468 | 0.848 (0.380 – 1.895) | 0.688 |
|  | Poor outcome | 2.137 (0.936 – 4.879) | 0.071 | 1.437 (0.615 – 3.354) | 0.402 |
| **GL02** | Severity | 3.082 (1.553 – 6.114) | 0.001<sup>b</sup> | 2.607 (1.278 – 5.320) | 0.008<sup>b</sup> |
|  | Poor outcome | 0.331 (0.078 – 1.416) | 0.136 | 0.255 (0.059 – 1.103) | 0.067 |
| **GL03** | Severity | 0.260 (0.062 – 1.093) | 0.066 | 0.526 (0.121 – 2.290) | 0.469 |
|  | Poor outcome | 0.179 (0.024 – 1.328) | 0.092 | 0.376 (0.049 – 2.862) | 0.345 |
| **GL04** | Severity | 0.279 (0.038 – 2.065) | 0.211 | 0.326 (0.043 – 2.480) | 0.279 |
|  | Poor outcome | 1.900 (0.638 – 5.664) | 0.249 | 2.476 (0.787 – 7.786) | 0.121 |
| **GL05** | Severity | 1.001 (0.233 – 4.297) | 0.999 | 1.314 (0.293 – 5.887) | 0.721 |
|  | Poor outcome | 1.431 (0.328 – 6.242) | 0.633 | 1.897 (0.416 – 8.642) | 0.408 |

<sup>a</sup> The numbers of patients for poor outcome were 862, as this information was missing for some patients.
<sup>b</sup> Benjamini-Hochberg-adjusted $P \leq 0.05$

### *emm* type STG62647 isolates

*emm* type STG62647 has emerged as the most prevalent cause of invasive SDSE infections in multiple surveillance studies in multiple countries over the past 15 years (14, 29–34). In some countries, this emergence has coincided with a perceived increase in the frequency and/or severity of infections leading to speculation that STG62647 isolates may be more virulent (19, 35). The 87 STG62647 isolates are the second most abundant *emm* type among the 865 isolates studied, accounting for 49% of GL01 isolates. Given the significant association between GL01 and skin/soft tissue infections, we examined whether STG62647 was also non-randomly associated with these infections. STG62647 was significantly over-represented among skin/soft tissue (*P* = 0.005), when tested in univariate analysis.

Given that both GL01 and STG62647 were individually associated with skin/soft tissue, this raised the question, is it genetic background or *emm* type that is most causally relevant? Only GL01 was significantly associated with skin/soft tissue infection (OR = 2.228, 95% CI (1.261–3.936), *P* = 0.006), whereas STG62647 was not independently associated (OR = 0.908, 95% CI (0.441–1.867), *P* = 0.793) in multivariate analysis. Importantly, further testing also found that STG62647 was not significantly associated with infection severity (OR = 0.541, 95% CI (0.219–1.340), *P* = 0.184) nor poor outcome (OR = 0.856, 95% CI (0.252–2.913), *P* = 0.804).

## DISCUSSION

We addressed the relative lack of genomic and patient epidemiologic studies of SDSE infections in the United States. Using WGS of 865 isolates cultured from consecutive patients in one large healthcare system in the Houston metroplex over two years, we identified significant genotype-phenotype associations. Integral to this molecular epidemiologic investigation was the availability of rich, comprehensive, and standardized clinical data collected from a single electronic medical record system, retrospectively and prospectively, which allowed us to study isolates causing both invasive and non-invasive infections. To our knowledge this study is the largest single collection of SDSE isolates of known clinical infection type characterized by whole genome sequencing. Infections caused by SDSE have been reported with increasing frequency in the last 10-15 years, and some investigators believe that the pathogen may represent an emerging infectious agent. Thus, analysis of a large comprehensive sample of SDSE isolates and the infections they cause is timely and was warranted.

Before the widespread use of WGS in molecular epidemiology, bacteria were commonly classified serologically on the basis of allelic variation in genes encoding one or a few cell-surface antigens, such as for the M protein or capsular polysaccharide. A limitation of such single genetic loci-based classifications is horizontal genetic transfer, resulting in serotype switching. Such switching events can be positively selected for by contributing to evasion of the host immune response and vaccines. This can compromise the utility of such classification schemes, especially in the case of species characterized by high levels of genomic diversity due to frequent episodes of horizontal gene transfer and recombination. Our congruence analyses comparing WGS data and different molecular classification schemes such as *emm* type or MLST confirmed that this is true for SDSE. That is, *emm* and MLST analysis are not sufficient for accurately capturing the phylogenetic structure of SDSE (14, 15, 19). In contrast to *S. pyogenes*, for which a strong association between *emm* types and GLs was determined (23), we found a poor correlation between *emm* type and less so MLST with phylogenetic population structure for the 865 SDSE isolates studied. Horizontal genetic transfer and recombination was extensive along the SDSE genome and in the phylogeny. Recombination was found nearly twice as abundant among SDSE housekeeping genes as in *S. pyogenes,* as determined with STs. These finding strongly supports the hypothesis that a high rate of HGT and recombination is the underlying molecular mechanism causing the low correlation between *emm* type and MLST with the population genomic structure in SDSE. Previous studies assessing recombination in SDSE reported a effective mutation rate comparable to *S. pyogenes* (19). A crucial difference resides in the absolute frequency and the magnitude of genetic diversity introduced by each recombination event. This suggests that although both species experience recombination, the genomic structure of SDSE is more rapidly diversified than for *S*. *pyogenes*. We and other investigators have found *S. pyogenes* to have a rate of recombination comparable to or exceeding that of naturally competent species, such as *Streptococcus pneumoniae* and *Neisseria meningitidis* (23, 24). However, effective recombination in *S. pyogenes* is consistent with transfer occurring over a restricted donor host range, that is primarily with another *S. pyogenes* or with SDSE. The greater diversity of recombination regions in SDSE than *S. pyogenes*, and the greater diversity present in the MLST housekeeping genes, suggests that SDSE likely has a broader donor host range for effective recombination.

Conclusively establishing isolate genotype-patient phenotype associations in SDSE has proven challenging, likely due to three primary limiting factors: 1) reliance on low-resolution genetic markers such as *emm* type and MLST, which fail to accurately capture the true population structure; 2) modest sample size, which lacks the statistical power required to overcome the high background diversity of the pathogen; and 3) absence of comprehensive, standardized clinical metadata necessary to accurately classify infection types and patient outcomes (18, 28, 36–38). Low-resolution genetic markers often obscure associations because they do not capture overall genomic relationships among isolates, leading to inconsistencies across global cohorts that reflect dependence on geographical setting and sample period, rather than inherent genomic differences (for a review see (10)). In our study, we addressed these limitations using a GL system based on genome-wide core SNP variation, coupled with analysis of a large, retrospectively and prospectively collected, comprehensive isolate sample (*n* = 865 isolates) for which we had high-quality matched patient information. This analysis strategy successfully identified significant associations between some SDSE GLs and clinical infection types. This discovery may pave the way to enhanced understanding of pathogen-host interaction at the gene or SNP level. It is reasonable to speculate that increasing sample size will yield additional information about associations, and studies are underway to test this idea.

The identification of pathologically relevant genotype-phenotype associations is aided by unbiased dense sampling with rich genetic and clinical information. When a clone becomes predominant such as for STG62647 (14, 34, 35, 39), it increases the likelihood that a coincidental association between clone and phenotype is misinterpreted as pathologically relevant. Additionally, ascertainment bias, such as the well-intentioned collection of more severe invasive infection isolates, can create spurious associations driven by sampling density rather than true biological virulence. This may be the case for STG62647, because only a modest number of SDSE isolates from limited time periods, geographic areas, and anatomic sites have been analyzed by WGS.

In our study, *emm* type STG62647 was not significantly associated with severe disease or poor infection outcome. This is consistent with the findings of Lopez De Egea et al (40) that STG62647 was not associated with severe clinical outcomes and Xie et al. (41) that there was no association with mortality rate. Furthermore, although prevalent in our study sample, STG62647 was not independently linked to skin/soft tissue infections in multivariate analysis. In contrast, GL01, to which all but one of STG62647 isolates belong, had a significant independent association with skin/soft tissue infection. Our data conclusively establish that GLs defined by genome-wide core SNPs variation, constitute the definitive marker for investigating SDSE genotype-patient phenotype associations. Given its speed and modest cost (comparable to characterization by *emm* and MLST sequencing), we recommend that subsequent SDSE epidemiological and clinical studies adopt whole genome sequencing, as analysis based on genetic classification using a single or only a few genes is demonstrably insufficient to accurately capture the population structure complexity of this highly recombinogenic pathogen.

The mean age of our patients with bacteremia (64 ± 15) is consistent with reports identifying an association between SDSE invasive infections and age > 60 years old (1, 8, 30, 32, 42–49). Similarly, our data confirm and expand previous observations linking invasive SDSE infections and high comorbidity burden, particularly cardiovascular disease and diabetes (31, 42, 49). Substantial demographic shifts are underway in the United States and most other high-income countries. Specifically, populations are aging (50) and cardiovascular diseases (51) and diabetes mellitus are more prevalent in high-income countries (52). This means that the number of individuals at risk for developing serious invasive infections caused by SDSE is increasing.

Thus, it is reasonable to think that SDSE infections will continue to be an important cause of serious disease in many countries and likely increase in frequency. These trends serve as an important impetus to further analyze SDSE molecular population genomics in defined populations. Similarly, increased investigative efforts are needed to identify molecular mechanisms contributing to SDSE-human interactions, including colonization, invasion, and host immune response.

Although this study provides the largest single SDSE whole-genome dataset characterized to date, it is subject to the inherent limitations of a single institutional collection. Analyses of SDSE isolated in other regions are required to place our findings in the context of a larger geographic sample. Secondly, the study only covered two years. This limited time frame was insufficient to perform robust evolutionary rate analyses. Thirdly, the absence of complete, closed genome sequences limits our ability to fully characterize quasi-repetitive mobile genetic elements that might carry crucial antimicrobial resistance genes and hinders the exploration of genomic structural variations with the potential to alter regulatory networks. In addition, little is known about what SDSE genes are expressed in patients, regardless of disease type. Future analyses such as transcriptome and animal infection model studies likely will help identify molecular mechanisms of pathogen-host interaction operative in SDSE human infections.

In conclusion, this study provides the largest population genomic analysis of SDSE isolates circulating in a large population center in the United States and thereby addresses a knowledge deficit for this emerging pathogen. Our findings highlight the need to use WGS and linked patient information for routine SDSE surveillance and molecular epidemiologic analysis to routinely investigate SDSE molecular population genomics, and infection type and severity.

## MATERIALS AND METHODS

### Study population

The study period was June 2022 to August 2024. The Houston Methodist (HM) hospital system has nine hospitals located in different geographic regions across the large metroplex, with an ethnically diverse population of 7.4 million. All specimens used for microbiology culture diagnostics are processed in one clinical laboratory. All isolates identified as *S. dysgalactiae* by matrix-assisted laser desorption ionization–time of flight mass spectrometry (MALDI-TOF MS) were saved and WGS was performed to determine subspecies and overall genomic relationships among SDSE isolates. Isolates were subcultured at 37°C with 5% CO_2_ on Columbia blood agar plates and stored at minus 80°C in Todd Hewitt Broth (THB, Legacy Biologicals, Baltimore, MD) supplemented with 0.2 % yeast extract and 20% glycerol. The study was approved by the Houston Methodist Institutional Review Board (IRB number PRO00001153).

### Identification of Lancefield group carbohydrate

Lancefield group carbohydrate antigen was determined by latex agglutination (BBL Streptocard enzyme latex test; Becton, Dickinson, Franklin Lakes, NJ) on overnight growth harvested from blood agar plates.

### Demographic and patient data

Patient demographic information such as age, gender, and ethnicity were obtained from the electronic medical record. Three age categories were used, including 1) pediatric patients, less than 18 years old; 2) adult patients, 18–64 years old; and 3) senior patients, 65 years old and older. Clinical information, such as diagnosis, presence of severe infection indicators (i.e., septic shock, ICU admission), poor outcomes (i.e., disability, amputation, death within 30 days of infection), and comorbidities (e.g., cardiovascular diseases, diabetes mellitus), were obtained by review of the electronic medical record, using a single electronic medical system (Epic).

### Exclusion of isolates cultured from the same patient

Multiple isolates cultured from the same patient were excluded from the analysis. Of the 996 consecutive SDSE isolates identified in the HM diagnostic microbiology laboratory during the study period, 131 isolates collected from same patient were excluded, resulting in a total of 865 unique SDSE isolates.

### Genome sequencing

SDSE genomes were sequenced as previously described (53). Briefly, isolates were grown at 37°C in 5% CO_2_ on tryptic soy agar with 5% sheep blood (Becton, Dickinson, Franklin Lakes, NJ). Chromosomal DNA for sequencing was isolated with the RNAdvance viral kit (Beckman Coulter, Brea, CA) automated on a BioMek i7 liquid handling robot (Beckman Coulter).

Illumina paired-end short read sequencing libraries were prepared with a NexteraXT kit (Illumina, San Diego, CA) and sequenced with a NovaSeq instrument using a 2 x 250-bp protocol.

### Genome assembly and analysis

Illumina sequencing reads were quality controlled and preprocessed with fastp (54) and assembled with SPAdes 3.9.0 (55). Genome assembly quality was assessed with QUAST (56). Taxonomic identification using genome assembly contigs were made with GAMBIT (57). *emm* types were determined with emmtyper (https://github.com/MDU-PHL/emmtyper) relative to the Center for Disease Control and Prevention *emm* type database (https://www2.cdc.gov/vaccines/biotech/strepblast.asp). Multilocus sequence types were determined with FastMLST (58) relative to the PubMLST database (https://pubmlst.org). Clonal complexes of STs, as defined by clustering of single locus variants, were determined using goeBURST (59). Polymorphisms relative to SDSE ST20/STG62647 reference strain MGCS36044 closed genome (GenBank accession GCA029234115.1) (53) were determined from sequencing reads with SNIPPY (https://github.com/tseemann/snippy), and from assembly contigs with MUMmer 4.0 (60). Phylogenies were inferred by neighbor-joining with RapidNJ (61) and by maximum-likelihood with RAxML-NG (62) and illustrated with Dendroscope (63).

### Population genomic structure

Structure with in the population based on the genetic distance between genome assemblies was determined with PopPUNK v2.7.0 (17) by the method of hierarchical density based spatial cluster of applications with noise (HDBSCAN). The initial variable length k-mer sketching was done with k-mer lengths from 13 to 29 nucleotides and a sketch size of 100,000. Model refinement was run on the initial HDBSCAN fit to improve the network score. Resultant clusters were designated genetic lineages. Mean genetic distances in terms of the mean number of core chromosomal SNPs, among isolates within a GL and between isolates of different GLs were determined using MEGA (64).

### Recombination analyses

Evolutionary rates of horizontal recombination and vertical single site mutation among all 865 SDSE genomes was inferred using ClonalFrameML (22). To limit this analysis to the core genome, SNPs were identified using SNIPPY relative to MGCS36044 core genome, that is the genome with the previously identified 9 regions of difference constituting the accessory genome excluded. The core genome SNPs for each of the 865 isolates were then individually transposed into the MGCS36044 core genome sequence using bcftools consensus, generating 865 pseudo-core genomes. A pseudo-core genome maximum likelihood phylogeny was inferred using RAxML-NG. The pseudo-core genome sequences and corresponding ML phylogeny were then analyzed with ClonalFrameML (22) to estimate global recombination parameters and Gubbins (65) to quantify recombination events within the phylogeny. This recombination analysis was analogously applied to GL01 to GL10 subsets of isolates to determine the extent of variation in recombination among the GLs. The variance of recombination metrics among the GLs was evaluated for statistically significant outliers using Grubb’s test. For recombination analysis of the 1,659 *Streptococcus pyogenes* and 824 *Streptococcus dysgalactiae* STs obtained from PubMLST, the ST sequences were aligned with MAFFT (66) prior to inferring ML phylogenies. This facilitated a direct comparison of the abundance of recombination events between the species.

### Statistical analysis

All statistical analyses were conducted with IBM SPSS Statistics version 21.0 (SPSS Inc., Chicago, IL, USA). For analysis of categorical data, we used primarily Pearson’s chi-square test. When the assumption of sufficient expected cell counts was violated (typically < 5 in any cell), we used Monte Carlo simulation of the chi-square test (with 10,000 simulated tables) or the Fisher-Freeman-Halton exact test. When chi-square test could not be performed due to small sample sizes (expected cell counts < 5), Fisher’s exact test was employed. Post-hoc analyses (pairwise comparisons) were conducted only when the global test of association was statistically significant. To mitigate the risk of Type I errors due to multiple comparisons, the resultant *P*-values were corrected with the Benjamini-Hochberg procedure to control the False Discovery Rate (FDR). These follow-up analyses were used to identify which category comparisons contributed most to the observed overall association. A two-sided significance threshold of *P* < 0.05 was applied for all statistical tests.

## Ethics statement

Patient information was obtained under Houston Methodist human subjects protocol PRO00001153.

## Data availability

Genome sequences have been submitted to the National Center for Biotechnology Information (NCBI) under Bioproject PRJNA1363881.

## SUPPLEMENTAL MATERIAL

Supplemental material is available online only.

**SUPPLEMENTAL Tables S1-S4**

**SUPPLEMENTAL FIG S1** SDSE-865 HDBSCAN refined fit

**SUPPLEMENTAL FIG S2.1** Recombination analysis GL01-GL05

**SUPPLEMENTAL FIG S2.2** Recombination analysis GL06-GL10

**SUPPLEMENTAL FIG S3** GL recombination metrics outlier assessment

## Supporting information

Table S1 Patient Demographic, Case Clinical, and Isolate Genetic Characteristics

Table S2 Patient Demographic and Case Clinical Associations

Table S3 Molecular Classification Schemes Comparisons

Table S4 GL Genotype and Infection Site Phenotype Associations

FIG S1 HDBCSAN Refined Fit

FIG S2.1 Recombination GL01-GL05

FIG S2.2 Recombination GL06-GL10

FIG S3 GL Recombination Metrics Grubb's Test

## ACKNOWLEDGMENTS

We declare no conflict of interest.

These studies were funded in part by the Fondren Foundation. We thank Lindsay Kowis, Patricia L Cernoch, Paul A Christensen, Matthew Ojeda Saavedra, Michele Byrom, Alma Amaya, Ryan Gadd, Regan Mangham, Eleanor Nichols, Jordan Pachuca, Sindy Pena, Kristina Reppond, Brenda Medreno, Catalina Mesa, Nada Sayed, and Andrea Torres Ramirez for technical assistance, and Sasha Pejerrey and Heather McConnell for editorial assistance.

---

**FIG S1** Illustrated is the relationship between pair-wise core and accessory genome distances for all 865 isolates of SDSE. The calculated decision boundary was used to partition the data into 44 distinct genetic lineages (GLs). The high network transitivity indicates good within clusters/GLs cohesion, and the low network density indicates that the population is highly structured into many discrete clusters/GLs.

**FIG S2** SDSE genetic lineage recombination assessment. Illustrated in panels A-to-L are phylogenetic trees (on the left) and recombination block diagrams (on the right) with a heatmap of recombination events (along the top) as inferred with Gubbins. Below each diagram are given recombination and mutation metrics for the core chromosomal sequences analyzed as determined with ClonalFrameML. The GL and number of isolates/core chromosomal sequences analyzed is given in the header of each panel.

**FIG S3** Genetic lineage recombination metrics outlier assessment. Illustrated as dot plots are the recombination metrics determined for the SDSE cohort and genetic lineages as determined for the core chromosome sequences with ClonalFrameML. The genetic lineage core chromosome sequences analyzed are color coded as given in the index. The recombination metric is indicated below each plot. The red bar indicates the mean, and the value of the mean and standard deviation are given above each plot. Grubb’s outlier test (i.e. Extreme Studentized Deviate test) was used to test if the metric with the greatest deviation from the mean was statistically significantly different from the others. None of the GLs had a most divergent recombination metric that was a significant outlier. Although recombination metrics varied among the GLs, the variation was not statistically significant, indicating that the GLs are evolving similarly in terms of recombination and single site mutation.

## REFERENCES

1. Broyles LN, Van Beneden C, Beall B, Facklam R, Shewmaker PL, Malpiedi P, Daily P, Reingold A, Farley MM. 2009. Population-Based Study of Invasive Disease Due to β-Hemolytic Streptococci of Groups Other than A and B. Clinical Infectious Diseases 48:706–712.

2. Facklam R. 2002. What Happened to the Streptococci: Overview of Taxonomic and Nomenclature Changes. Clinical Microbiology Reviews 15:613–630.

3. Brandt CM, Spellerberg B. 2009. Human infections due to Streptococcus dysgalactiae subspecies equisimilis. Clinical infectious diseases: an official publication of the Infectious Diseases Society of America 49:766–772.

4. Bruun T, Rath E, Madsen MB, Oppegaard O, Nekludov M, Arnell P, Karlsson Y, Babbar A, Bergey F, Itzek A, Hyldegaard O, Norrby-Teglund A, Skrede S. 2021. Risk Factors and Predictors of Mortality in Streptococcal Necrotizing Soft-tissue Infections: A Multicenter Prospective Study. Clinical infectious diseases: an official publication of the Infectious Diseases Society of America 72:293–300.

5. Fuursted K, Stegger M, Hoffmann S, Lambertsen L, Andersen PS, Deleuran M, Thomsen MK. 2016. Description and characterization of a penicillin-resistant Streptococcus dysgalactiae subsp. equisimilis clone isolated from blood in three epidemiologically linked patients. Journal of Antimicrobial Chemotherapy 71:3376–3380.

6. Hagiya H, Okita S, Kuroe Y, Nojima H, Otani S, Sugiyama J, Naito H, Kawanishi S, Hagioka S, Morimoto N. 2013. A fatal case of streptococcal toxic shock syndrome due to Streptococcus dysgalactiae subsp. equisimilis possibly caused by an intramuscular injection. Internal medicine (Tokyo, Japan) 52:397–402.

7. Haidan A, Talay SR, Rohde M, Sriprakash KS, Currie BJ, Chhatwal GS. 2000. Pharyngeal carriage of group C and group G streptococci and acute rheumatic fever in an Aboriginal population. The Lancet 356:1167–1169.

8. Nevanlinna V, Huttunen R, Aittoniemi J, Luukkaala T, Rantala S. 2023. Incidence, seasonal pattern, and clinical manifestations of Streptococcus dysgalactiae subspecies equisimilis bacteremia; a population-based study. European journal of clinical microbiology & infectious diseases: official publication of the European Society of Clinical Microbiology 42:819–825.

9. Rantala S, Vahakuopus S, Vuopio-Varkila J, Vuento R, Syrjanen J. 2010. Streptococcus dysgalactiae subsp. equisimilis Bacteremia, Finland, 1995-2004. Emerging infectious diseases 16:843-846.

10. Xie O, Davies MR, Tong SYC. 2024. Streptococcus dysgalactiae subsp. equisimilis infection and its intersection with Streptococcus pyogenes. Clinical microbiology reviews 37:e0017523.

11. Bisno AL, Craven DE, McCabe WR. 1987. M proteins of group G streptococci isolated from bacteremic human infections. Infect Immun 55:753–7.

12. Collins CM, Kimura A, Bisno AL. 1992. Group G streptococcal M protein exhibits structural features analogous to those of class I M protein of group A streptococci. Infect Immun 60:3689–96.

13. Ahmad Y, Gertz RE, Jr., Li Z, Sakota V, Broyles LN, Van Beneden C, Facklam R, Shewmaker PL, Reingold A, Farley MM, Beall BW. 2009. Genetic relationships deduced from emm and multilocus sequence typing of invasive Streptococcus dysgalactiae subsp. equisimilis and S. canis recovered from isolates collected in the United States. J Clin Microbiol 47:2046–54.

14. Kaci A, Jonassen CM, Skrede S, Sivertsen A, Steinbakk M, Oppegaard O. 2023. Genomic epidemiology of Streptococcus dysgalactiae subsp. equisimilis strains causing invasive disease in Norway during 2018. Frontiers in microbiology 14:1171913.

15. McMillan DJ, Bessen DE, Pinho M, Ford C, Hall GS, Melo-Cristino J, Ramirez M. 2010. Population genetics of Streptococcus dysgalactiae subspecies equisimilis reveals widely dispersed clones and extensive recombination. PloS one 5:e11741.

16. Ruth D. 2012. Houston region Grows more ethnically diverse, with small declines in segrgation. Rice University Kinder Institute and Hobby Center for the Study of Texas.

17. Lees JA, Harris SR, Tonkin-Hill G, Gladstone RA, Lo SW, Weiser JN, Corander J, Bentley SD, Croucher NJ. 2019. Fast and flexible bacterial genomic epidemiology with PopPUNK. Genome research 29:304–316.

18. Pinho MD, Melo-Cristino J, Ramirez M. 2006. Clonal relationships between invasive and noninvasive Lancefield group C and G streptococci and emm-specific differences in invasiveness. J Clin Microbiol 44:841–6.

19. Xie O, Morris JM, Hayes AJ, Towers RJ, Jespersen MG, Lees JA, Ben Zakour NL, Berking O, Baines SL, Carter GP, Tonkin-Hill G, Schrieber L, McIntyre L, Lacey JA, James TB, Sriprakash KS, Beatson SA, Hasegawa T, Giffard P, Steer AC, Batzloff MR, Beall BW, Pinho MD, Ramirez M, Bessen DE, Dougan G, Bentley SD, Walker MJ, Currie BJ, Tong SYC, McMillan DJ, Davies MR. 2024. Inter-species gene flow drives ongoing evolution of Streptococcus pyogenes and Streptococcus dysgalactiae subsp. equisimilis. Nature Communications 15:2286.

20. McNeilly CL, McMillan DJ. 2014. Horizontal gene transfer and recombination in Streptococcus dysgalactiae subsp. equisimilis. Front Microbiol 5:676.

21. Croucher NJ, Page AJ, Connor TR, Delaney AJ, Keane JA, Bentley SD, Parkhill J, Harris SR. 2015. Rapid phylogenetic analysis of large samples of recombinant bacterial whole genome sequences using Gubbins. Nucleic Acids Research 43:e15–e15.

22. Didelot X, Wilson DJ. 2015. ClonalFrameML: efficient inference of recombination in whole bacterial genomes. PLoS computational biology 11:e1004041.

23. Beres SB, Zhu L, Pruitt L, Olsen RJ, Faili A, Kayal S, Musser JM. 2022. Integrative Reverse Genetic Analysis Identifies Polymorphisms Contributing to Decreased Antimicrobial Agent Susceptibility in Streptococcus pyogenes. mBio 13:e0361821.

24. Didelot X, Maiden MC. 2010. Impact of recombination on bacterial evolution. Trends Microbiol 18:315–22.

25. Duchene S, Holt KE, Weill FX, Le Hello S, Hawkey J, Edwards DJ, Fourment M, Holmes EC. 2016. Genome-scale rates of evolutionary change in bacteria. Microb Genom 2:e000094.

26. Gibson B, Eyre-Walker A. 2019. Investigating Evolutionary Rate Variation in Bacteria. J Mol Evol 87:317–326.

27. Hanage WP. 2016. Not So Simple After All: Bacteria, Their Population Genetics, and Recombination. Cold Spring Harb Perspect Biol 8.

28. Davies MR, McMillan DJ, Beiko RG, Barroso V, Geffers R, Sriprakash KS, Chhatwal GS. 2007. Virulence profiling of Streptococcus dysgalactiae subspecies equisimilis isolated from infected humans reveals 2 distinct genetic lineages that do not segregate with their phenotypes or propensity to cause diseases. Clinical infectious diseases: an official publication of the Infectious Diseases Society of America 44:1442–1454.

29. Itzek A, Weißbach V, Meintrup D, Rieß B, van der Linden M, Borgmann S. 2023. Epidemiological and Clinical Features of Streptococcus dysgalactiae ssp. equisimilis stG62647 and Other emm Types in Germany. Pathogens 12.

30. Lambertsen LM, Ingels H, Schønheyder HC, Hoffmann S. 2014. Nationwide laboratory-based surveillance of invasive beta-haemolytic streptococci in Denmark from 2005 to 2011. Clinical Microbiology and Infection 20:O216–O223.

31. Lother SA, Demczuk W, Martin I, Mulvey M, Dufault B, Lagacé-Wiens P, Keynan Y. 2017. Clonal Clusters and Virulence Factors of Group C and G Streptococcus Causing Severe Infections, Manitoba, Canada, 2012-2014. Emerging infectious diseases 23:1079–1088.

32. Rojo-Bezares B, Toca L, Azcona-Gutiérrez JM, Ortega-Unanue N, Toledano P, Sáenz Y. 2021. Streptococcus dysgalactiae subsp. equisimilis from invasive and non-invasive infections in Spain: combining epidemiology, molecular characterization, and genetic diversity. European journal of clinical microbiology & infectious diseases: official publication of the European Society of Clinical Microbiology 40:1013–1021.

33. Rossler S, Berner R, Jacobs E, Toepfner N. 2018. Prevalence and molecular diversity of invasive Streptococcus dysgalactiae and Streptococcus pyogenes in a German tertiary care medical centre. Eur J Clin Microbiol Infect Dis 37:1325–1332.

34. Trell K, Sendi P, Rasmussen M. 2016. Recurrent bacteremia with Streptococcus dysgalactiae: a case-control study. Diagnostic microbiology and infectious disease 85:121–124.

35. Oppegaard O, Mylvaganam H, Skrede S, Lindemann PC, Kittang BR. 2017. Emergence of a Streptococcus dysgalactiae subspecies equisimilis stG62647-lineage associated with severe clinical manifestations. Scientific Reports 7:7589.

36. Halperin T, Levine H, Korenman Z, Burstein S, Amber R, Sela T, Valinsky L. 2016. Molecular characterization and antibiotic resistance of group G streptococci in Israel: comparison of invasive, non-invasive and carriage isolates. European journal of clinical microbiology & infectious diseases: official publication of the European Society of Clinical Microbiology 35:1649–1654.

37. Jensen A, Kilian M. 2012. Delineation of Streptococcus dysgalactiae, its subspecies, and its clinical and phylogenetic relationship to Streptococcus pyogenes. J Clin Microbiol 50:113–26.

38. Leitner E, Zollner-Schwetz I, Zarfel G, Masoud-Landgraf L, Gehrer M, Wagner-Eibel U, Grisold AJ, Feierl G. 2015. Prevalence of emm types and antimicrobial susceptibility of Streptococcus dysgalactiae subsp. equisimilis in Austria. International journal of medical microbiology: IJMM 305:918–924.

39. Oppegaard O, Glambek M, Skutlaberg DH, Skrede S, Sivertsen A, Kittang BR. 2023. Streptococcus dysgalactiae Bloodstream Infections, Norway, 1999-2021. Emerging infectious diseases 29:260–267.

40. Lopez de Egea G, Gonzalez-Diaz A, Olsen RJ, Guedon G, Berbel D, Grau I, Camara J, Saiz-Escobedo L, Calvo-Silveria S, Cadenas-Jimenez I, Marimon JM, Cercenado E, Casabella A, Marti S, Dominguez MA, Leblond-Bourget N, Musser JM, Ardanuy C. 2025. Emergence of invasive Streptococcus dysgalactiae subsp. equisimilis in Spain (2012-2022): genomic insights and clinical correlations. Int J Infect Dis 153:107778.

41. Xie O, Featherstone L, Nguyen ANT, Hayes AJ, Pitt ME, Spring S, Liu A, Tonkin-Hill G, Dotel R, Joshi Rai N, Rofe A, Duchene S, Holt DC, Judd LM, Coin LJM, Krause VL, O’Sullivan MVN, Baird RW, Bond K, Howden BP, Korman TM, Currie BJ, Davies MR, Tong SYC. 2025. Invasive Streptococcus dysgalactiae subspecies equisimilis compared with Streptococcus pyogenes in Australia, 2011-23, and the emergence of a multi-continent stG62647 lineage: a retrospective clinical and genomic epidemiology study. Lancet Microbe 6:101182.

42. Couture-Cossette A, Carignan A, Mercier A, Desruisseaux C, Valiquette L, Pépin J. 2018. Secular trends in incidence of invasive beta-hemolytic streptococci and efficacy of adjunctive therapy in Quebec, Canada, 1996-2016. PloS one 13:e0206289.

43. Hanada S, Wajima T, Takata M, Morozumi M, Shoji M, Iwata S, Ubukata K. 2024. Clinical manifestations and biomarkers to predict mortality risk in adults with invasive Streptococcus dysgalactiae subsp. equisimilis infections. European journal of clinical microbiology & infectious diseases: official publication of the European Society of Clinical Microbiology 43:1609–1619.

44. Jaalama M, Palomäki O, Vuento R, Jokinen A, Uotila J. 2018. Prevalence and Clinical Significance of Streptococcus dysgalactiae subspecies equisimilis (Groups C or G Streptococci) Colonization in Pregnant Women: A Retrospective Cohort Study. Infectious diseases in obstetrics and gynecology 2018:2321046.

45. Liao C-H, Liu L-C, Huang Y-T, Teng L-J, Hsueh P-R. 2008. Bacteremia caused by group G Streptococci, taiwan. Emerging infectious diseases 14:837–840.

46. Solanki P, Colaco C, Dotel R. 2024. Analysis of bacteraemia caused by group C and G Streptococcus (Streptococcus dysgalactiae subsp. equisimilis) in Western Sydney over a 6-year period (2015-2020). European journal of clinical microbiology & infectious diseases: official publication of the European Society of Clinical Microbiology 43:1807–1814.

47. Takahashi T, Sunaoshi K, Sunakawa K, Fujishima S, Watanabe H, Ubukata K. 2010. Clinical aspects of invasive infections with Streptococcus dysgalactiae ssp. equisimilis in Japan: differences with respect to Streptococcus pyogenes and Streptococcus agalactiae infections. Clinical microbiology and infection: the official publication of the European Society of Clinical Microbiology and Infectious Diseases 16:1097–1103.

48. Tsuchihashi Y, Tamura K, Matsumoto K, Mitsushima S, Fujiya Y, Kuronuma K, Tanabe Y, Kasahara K, Maruyama T, Gotoh K, Nakamatsu M, Oshima K, Abe S, Nishi J, Arakawa Y, Ikebe T, Sunagawa T, Akeda Y, Oishi K. 2025. Comparative analysis of streptococcal toxic shock syndrome caused by three β-hemolytic streptococcal species in Japan. International journal of infectious diseases: IJID: official publication of the International Society for Infectious Diseases 158:107962.

49. Wajima T, Morozumi M, Hanada S, Sunaoshi K, Chiba N, Iwata S, Ubukata K. 2016. Molecular Characterization of Invasive Streptococcus dysgalactiae subsp. equisimilis, Japan. Emerging infectious diseases 22:247–254.

50. Bureau USC. May 18 2023. 2020 Census: United States Older Population Grew. https://www.census.gov/library/stories/2023/05/2020-census-united-states-older-population-grew.html. Accessed 11/10/2025.

51. Lindstrom M, DeCleene N, Dorsey H, Fuster V, Johnson CO, LeGrand KE, Mensah GA, Razo C, Stark B, Varieur Turco J, Roth GA. 2022. Global Burden of Cardiovascular Diseases and Risks Collaboration, 1990-2021. Journal of the American College of Cardiology 80:2372–2425.

52. Collaborators GBDD. 2023. Global, regional, and national burden of diabetes from 1990 to 2021, with projections of prevalence to 2050: a systematic analysis for the Global Burden of Disease Study 2021. Lancet 402:203–234.

53. Beres SB, Olsen RJ, Long SW, Eraso JM, Boukthir S, Faili A, Kayal S, Musser JM. 2023. Analysis of the Genomics and Mouse Virulence of an Emergent Clone of Streptococcus dysgalactiae Subspecies equisimilis. Microbiology spectrum 11:e0455022.

54. Chen S. 2023. Ultrafast one-pass FASTQ data preprocessing, quality control, and deduplication using fastp. iMeta 2:e107.

55. Bankevich A, Nurk S, Antipov D, Gurevich AA, Dvorkin M, Kulikov AS, Lesin VM, Nikolenko SI, Pham S, Prjibelski AD, Pyshkin AV, Sirotkin AV, Vyahhi N, Tesler G, Alekseyev MA, Pevzner PA. 2012. SPAdes: a new genome assembly algorithm and its applications to single-cell sequencing. J Comput Biol 19:455–77.

56. Gurevich A, Saveliev V, Vyahhi N, Tesler G. 2013. QUAST: quality assessment tool for genome assemblies. Bioinformatics (Oxford, England) 29:1072–1075.

57. Lumpe J, Gumbleton L, Gorzalski A, Libuit K, Varghese V, Lloyd T, Tadros F, Arsimendi T, Wagner E, Stephens C, Sevinsky J, Hess D, Pandori M. 2023. GAMBIT (Genomic Approximation Method for Bacterial Identification and Tracking): A methodology to rapidly leverage whole genome sequencing of bacterial isolates for clinical identification. PLoS One 18:e0277575.

58. Guerrero-Araya E, Muñoz M, Rodríguez C, Paredes-Sabja D. 2021. FastMLST: A Multi-core Tool for Multilocus Sequence Typing of Draft Genome Assemblies. Bioinformatics and biology insights 15:11779322211059238.

59. Francisco AP, Bugalho M, Ramirez M, Carrico JA. 2009. Global optimal eBURST analysis of multilocus typing data using a graphic matroid approach. BMC Bioinformatics 10:152.

60. Marçais G, Delcher AL, Phillippy AM, Coston R, Salzberg SL, Zimin A. 2018. MUMmer4: A fast and versatile genome alignment system. PLoS computational biology 14:e1005944.

61. M Simonsen TM, CNS Pedersen. 2008. Rapid neighbour-joining, p 113-122. In Lagergren KCJ (ed), Algorithms in Bioinformatics. Springer Berlin Heidelberg, Berlin, Heidelberg.

62. Kozlov AM, Darriba D, Flouri T, Morel B, Stamatakis A. 2019. RAxML-NG: a fast, scalable and user-friendly tool for maximum likelihood phylogenetic inference. Bioinformatics 35:4453–4455.

63. Huson DH, Scornavacca C. 2012. Dendroscope 3: an interactive tool for rooted phylogenetic trees and networks. Syst Biol 61:1061–7.

64. Kumar S, Stecher G, Suleski M, Sanderford M, Sharma S, Tamura K. 2024. MEGA12: Molecular Evolutionary Genetic Analysis Version 12 for Adaptive and Green Computing. Mol Biol Evol 41.

65. Croucher NJ, Didelot X. 2015. The application of genomics to tracing bacterial pathogen transmission. Current opinion in microbiology 23:62–67.

66. Katoh K, Misawa K, Kuma K, Miyata T. 2002. MAFFT: a novel method for rapid multiple sequence alignment based on fast Fourier transform. Nucleic Acids Res 30:3059–66.

