## Supplementary material for "Genomic and Patient Epidemiology of *Streptococcus dysgalactiae* Subspecies *equisimilis* in Houston, Texas": FIG S1 HDBCSAN Refined Fit

**Supplemental FIG S1 SDSE-865 PopPUNK HDBSCAN Refined Fit**

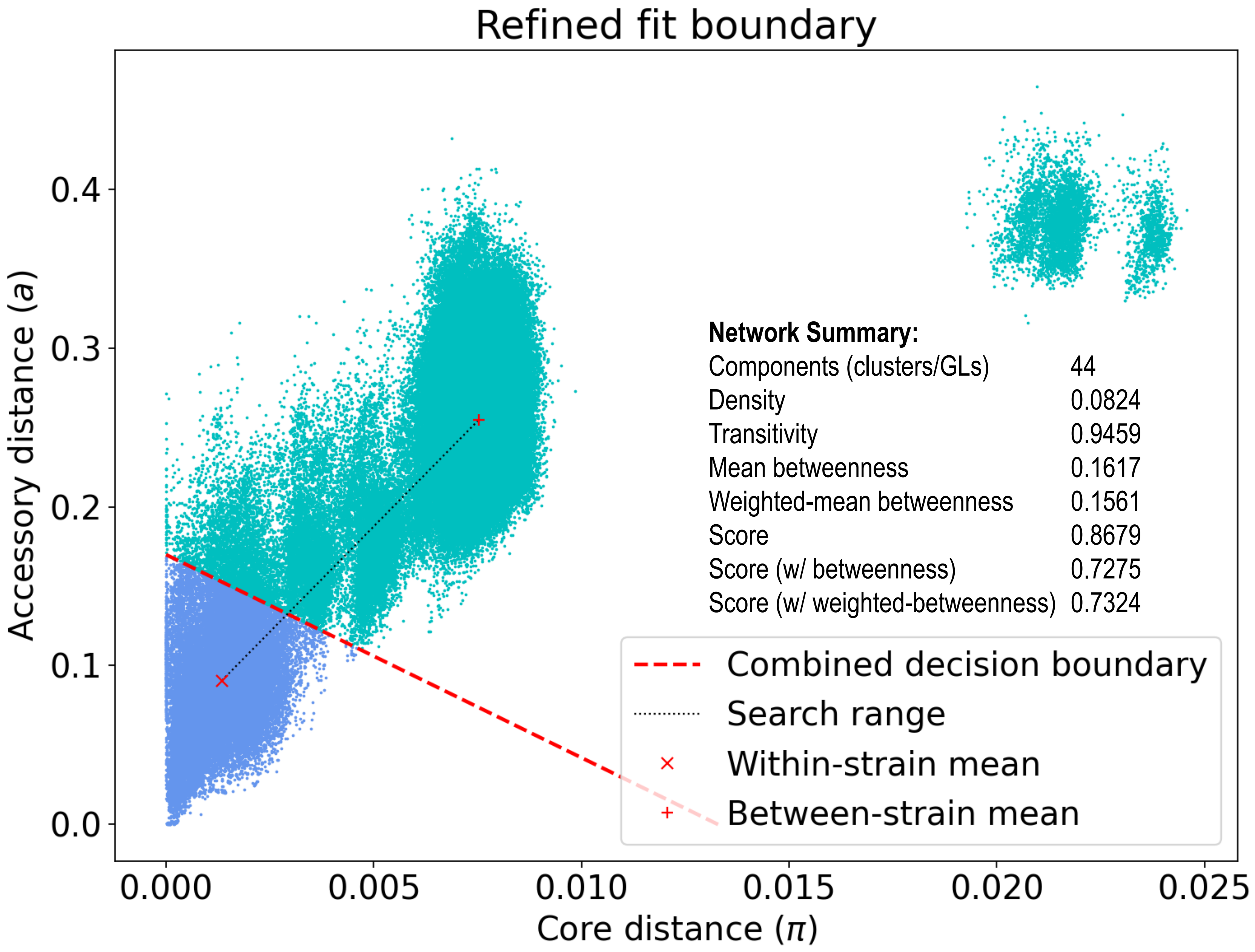


Illustrated is the relationship between pair-wise core and accessory genome distances for all 865 isolates of SDSE. The calculated decision boundary was used to partition the data into 44 distinct Genetic Lineages (GLs). The high network transitivity indicates good within clusters/GLs cohesion, and the low network density indicates that the population is highly structured into many discrete clusters/GLs
