## Supplementary material for "Genomic and Patient Epidemiology of *Streptococcus dysgalactiae* Subspecies *equisimilis* in Houston, Texas": FIG S2.1 Recombination GL01-GL05

### Supplemental FIG S2.1 SDSE genetic lineage recombination assessment

**A** **HMH SDSE Cohort, *n* = 865 Isolates**

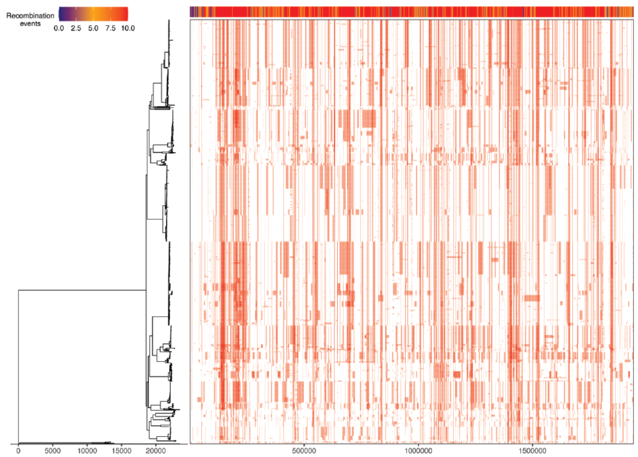


$\rho/\theta$ = 0.3451
$\delta$ = 129.87
$\nu$ = 0.0541
$\delta\nu$ = 7.030
$\rho\delta\nu/\theta$ = 2.4260

**B** **Genetic Lineage GL01, *n* = 175 Isolates**

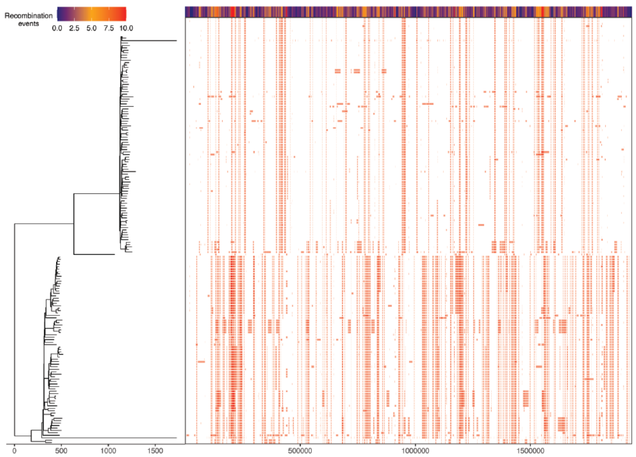


$\rho/\theta$ = 0.3278
$\delta$ = 152.56
$\nu$ = 0.0486
$\delta\nu$ = 7.428
$\rho\delta\nu/\theta$ = 2.4356

**C** **Genetic Lineage GL02, *n* = 171 Isolates**

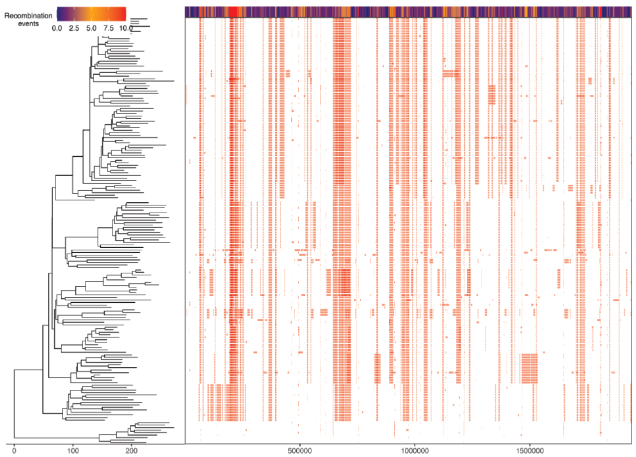


$\rho/\theta$ = 0.4363
$\delta$ = 191.98
$\nu$ = 0.0352
$\delta\nu$ = 6.762
$\rho\delta\nu/\theta$ = 2.9509

**D** **Genetic Lineage GL03, *n* = 155 Isolates**

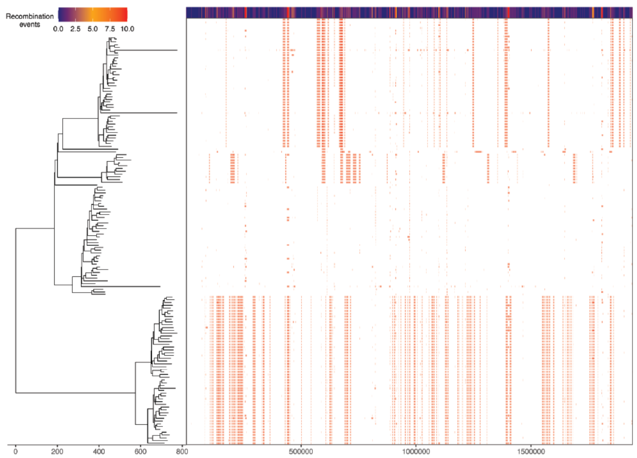


$\rho/\theta$ = 0.3639
$\delta$ = 164.44
$\nu$ = 0.0372
$\delta\nu$ = 6.134
$\rho\delta\nu/\theta$ = 2.2320

**E** **Genetic Lineage GL04, *n* = 78 Isolates**

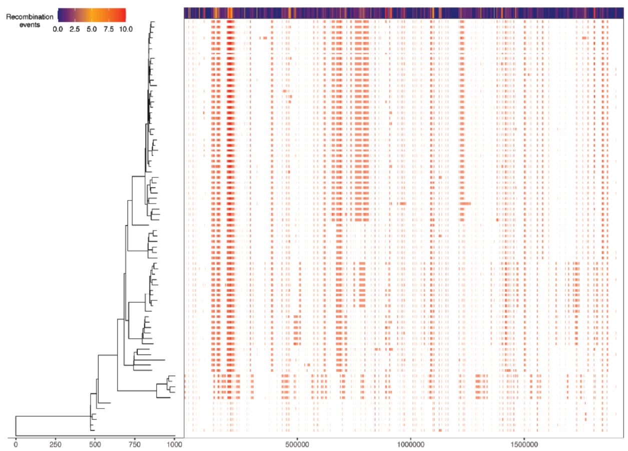


$\rho/\theta$ = 0.5728
$\delta$ = 217.77
$\nu$ = 0.0291
$\delta\nu$ = 6.333
$\rho\delta\nu/\theta$ = 3.6279

**F** **Genetic Lineage GL05, *n* = 48 Isolates**

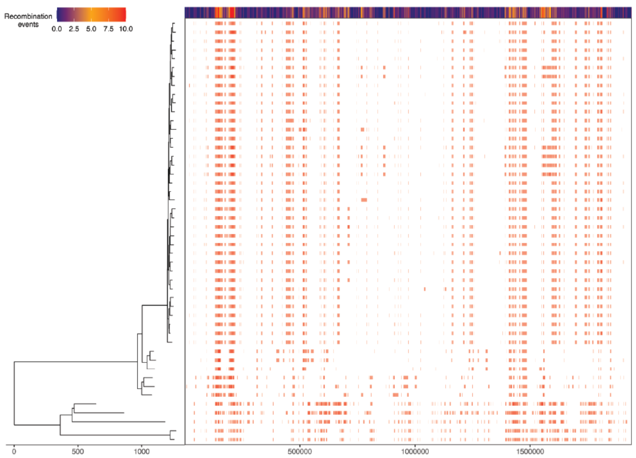


$\rho/\theta$ = 0.5234
$\delta$ = 200.89
$\nu$ = 0.0384
$\delta\nu$ = 7.707
$\rho\delta\nu/\theta$ = 4.0337

Illustrated in panels A-to-L are phylogenetic trees (on the left) and recombination block diagrams (on the right) with a heatmap of recombination events (along the top) as inferred with Gubbins. Below each diagram are given recombination and mutation metrics for the core chromosomal sequences analyzed as determined with ClonalFrameML. The GL and number of isolates/core chromosomal sequences analyzed is given in the header of each panel.
