## Supplementary material for "Genomic and Patient Epidemiology of *Streptococcus dysgalactiae* Subspecies *equisimilis* in Houston, Texas": FIG S2.2 Recombination GL06-GL10

### Supplemental FIG S2.2. SDSE genetic lineage recombination assessment

#### G Genetic Lineage GL06, *n* = 44 Isolates

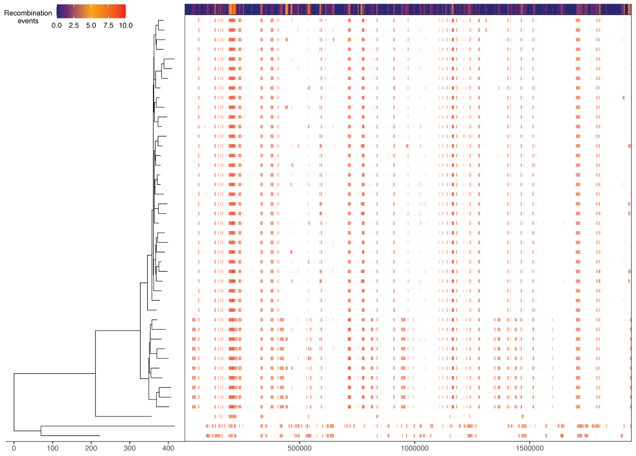


$\rho/\theta$ = 0.4552
$\delta$ = 234.24
$\nu$ = 0.0363
$\delta\nu$ = 8.498
$\rho\delta\nu/\theta$ = 3.8680

#### H Genetic Lineage GL07, *n* = 36 Isolates

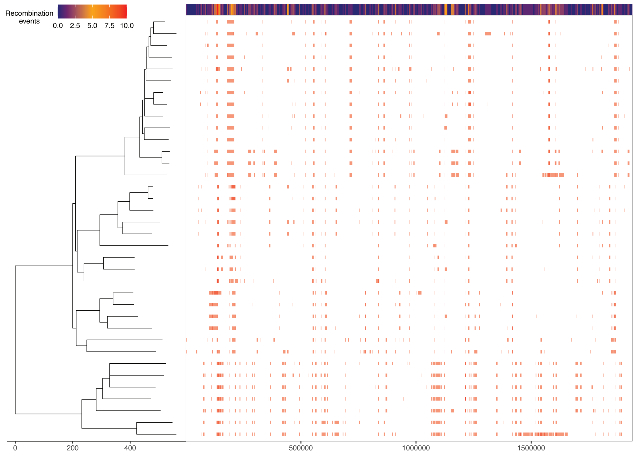


$\rho/\theta$ = 0.2804
$\delta$ = 213.03
$\nu$ = 0.0471
$\delta\nu$ = 10.024
$\rho\delta\nu/\theta$ = 2.8103

#### I Genetic Lineage GL08, *n* = 26 Isolates

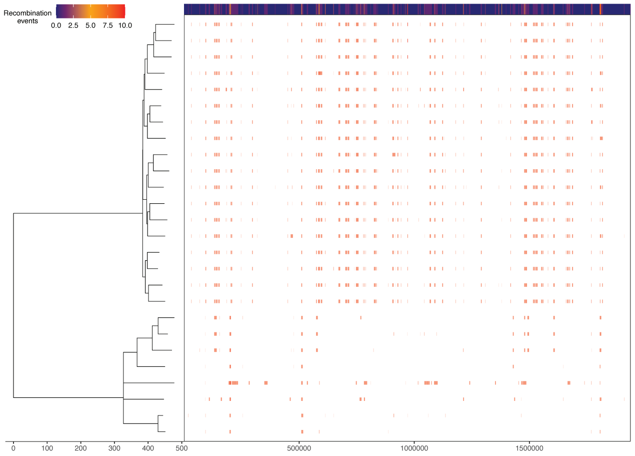


$\rho/\theta$ = 0.3505
$\delta$ = 172.87
$\nu$ = 0.0470
$\delta\nu$ = 8.118
$\rho\delta\nu/\theta$ = 2.8455

#### J Genetic Lineage GL09, *n* = 22 Isolates

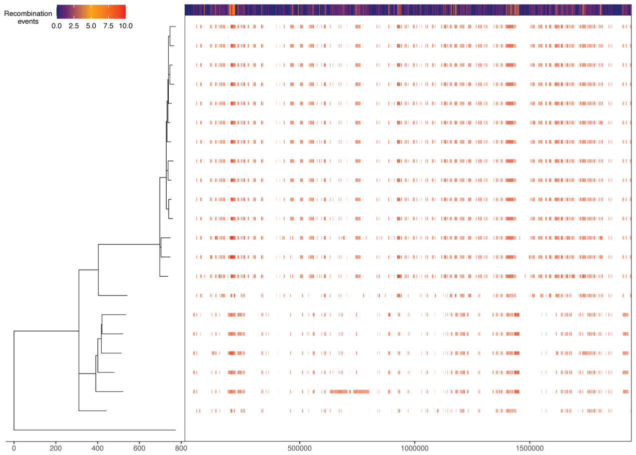


$\rho/\theta$ = 0.4267
$\delta$= 194.47
$\nu$ = 0.0461
$\delta\nu$ = 8.959
$\rho\delta\nu/\theta$= 3.8230

#### K Genetic Lineage GL10, *n* = 20 Isolates

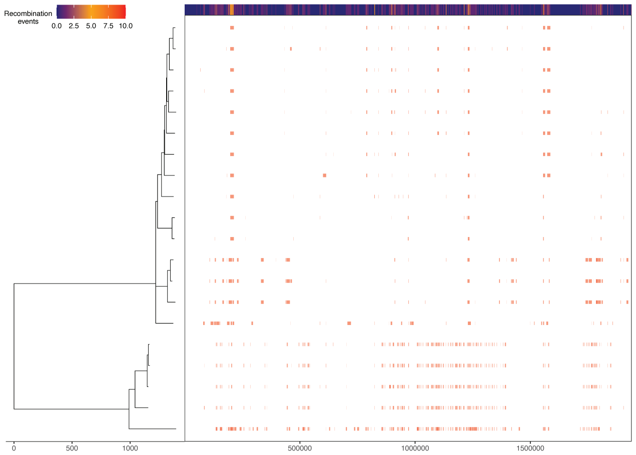


$\rho/\theta$ = 0.3656
$\delta$ = 196.88
$\nu$ = 0.0490
$\delta\nu$ = 9.647
$\rho\delta\nu/\theta$ = 3.5272

#### L Genetic Lineages GL11-to-GL44, *n* = 90 Isolates

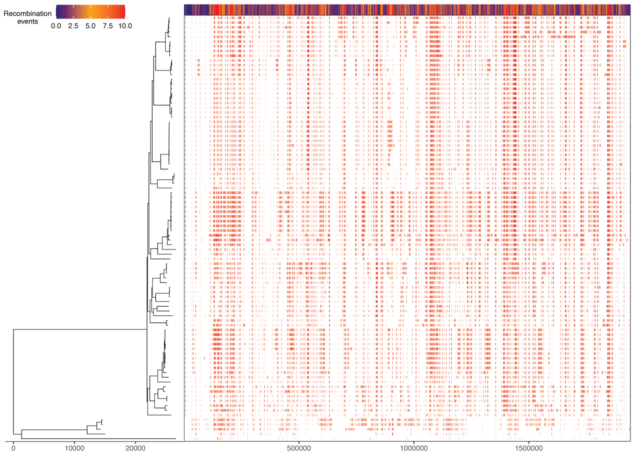


$\rho/\theta$ = 0.2856
$\delta$ = 95.72
$\nu$ = 0.0712
$\delta\nu$ = 6.819
$\rho\delta\nu/\theta$ = 1.9479

Illustrated in panels A-to-L are phylogenetic trees (on the left) and recombination block diagrams (on the right) with a heatmap of recombination events (along the top) as inferred with Gubbins. Below each diagram are given recombination and mutation metrics for the core chromosomal sequences analyzed as determined with ClonalFrameML. The GL and number of isolates/core chromosomal sequences analyzed is given in the header of each panel.
