## Supplementary material for "Genomic and Patient Epidemiology of *Streptococcus dysgalactiae* Subspecies *equisimilis* in Houston, Texas": FIG S3 GL Recombination Metrics Grubb's Test

### Supplemental FIG S3 Genetic lineage recombination metrics outlier assessment

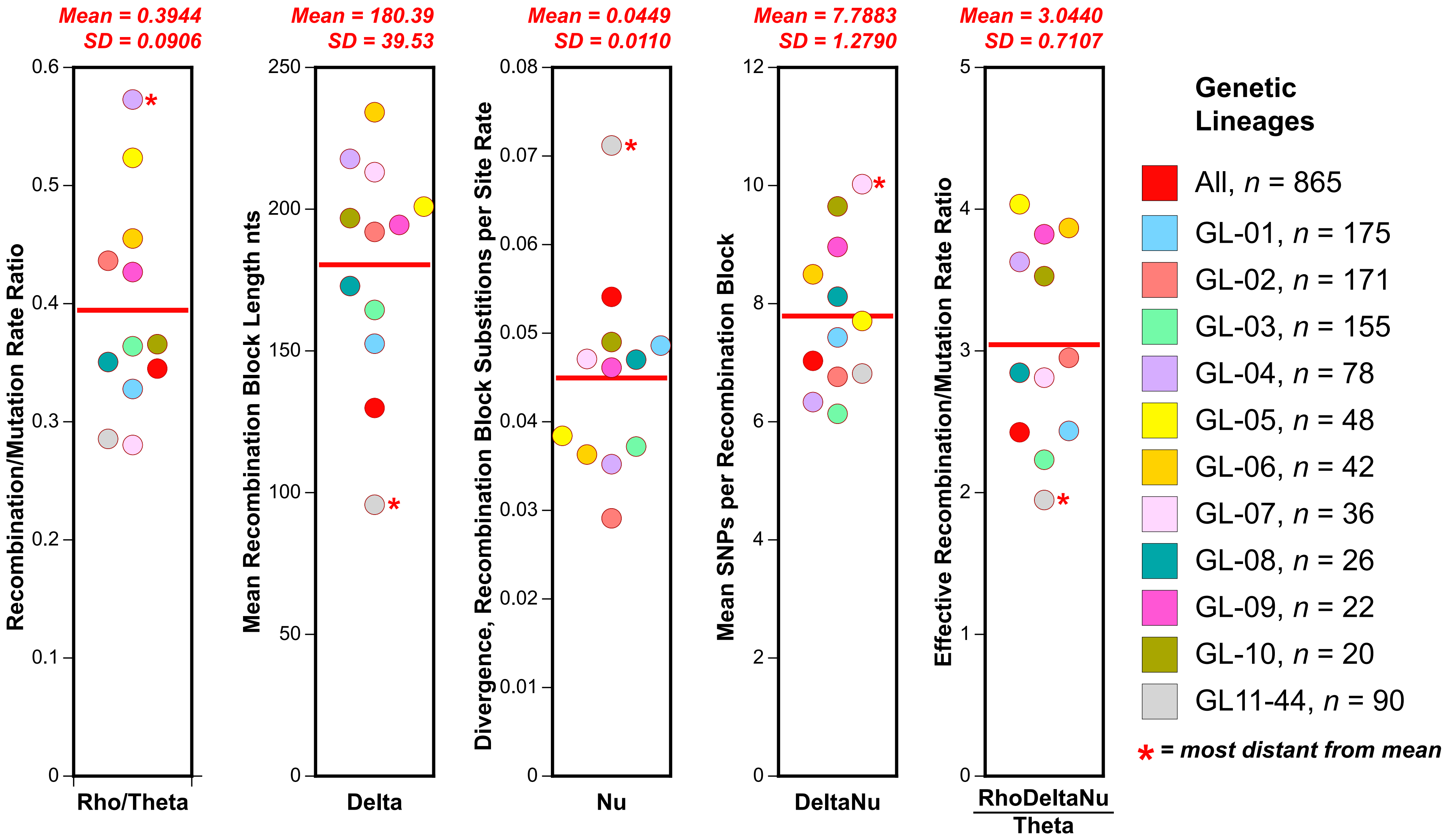


Illustrated as dot plots are the recombination metrics determined for the SDSE cohort and genetic lineages as determined for the core chromosome sequences with ClonalFrameML. The genetic lineage core chromosome sequences analyzed are color coded as given in the index. The recombination metric is indicated below each plot. The red bar indicates the mean, and the value of the mean and standard deviation are given above each plot. Grubb's outlier test (i.e. Extreme Studentized Deviate test) was used to test if the metric with the greatest deviation from the mean was statistically significantly different from the others. None of the GLs had a most divergent recombination metric that was a significant outlier. Although recombination metrics varied among the GLs, the variation was not statistically significant, indicating that the GLs are evolving similarly in terms of recombination and single site mutation.
